# Mediators of Treatment Response in Clinical Trial of Naltrexone and Bupropion for Methamphetamine Use Disorder: A Longitudinal Mediation Analysis

**DOI:** 10.64898/2026.05.09.26352807

**Authors:** Ramin Mojtabai, Ryoko Susukida, Mehdi Farokhnia, Trang Quynh Nguyen, Lorenzo Leggio, Cecilia Bergeria, Smita Prasad, Kelly E. Dunn, Masoumeh Amin-Esmaeili

**Affiliations:** Department of Psychiatry and Behavioral Sciences, Tulane Medical School, New Orleans, LA, USA; Department of Mental Health, Johns Hopkins Bloomberg School of Public Health, Baltimore, MD, USA; Clinical Psychoneuroendocrinology and Neuropsychopharmacology Section, Translational Addiction Medicine Branch, National Institute on Drug Abuse Intramural Research Program and National Institute on Alcohol Abuse and Alcoholism Division of Intramural Clinical and Biological Research, National Institutes of Health, Baltimore and Bethesda, MD, USA; Kahlert Institute for Addiction Medicine, University of Maryland School of Medicine, Baltimore MD, USA

**Keywords:** methamphetamine, stimulants, mediators, treatment effect mechanisms, secondary data analysis, craving, multilevel structural equation modeling

## Abstract

**Background:** The mechanisms underlying pharmacological treatments for stimulant use disorders are poorly understood. This study examined whether changes in craving, depressive symptoms, and/or impulsivity mediate treatment effect in pharmacotherapy with combined naltrexone and bupropion for methamphetamine use disorder.

**Methods:** The study was based on secondary analysis of data from the Accelerated Development of Additive Pharmacotherapy Treatment for methamphetamine disorder (ADAPT-2) trial which randomized adults with methamphetamine use disorder to combined treatment with injectable naltrexone (380 mg every three weeks) plus oral bupropion (450 mg daily) versus placebo. A total of 403 adults with methamphetamine use disorder participated in the first Stage; 225 of first Stage participants in the placebo arm who did not respond to treatment were re-randomized in the second Stage. Mediation effects were examined using longitudinal multi-level structural equation modeling.

**Results:** Naltrexone-bupropion treatment was associated with decreases in drug use, craving, depressive symptoms, and impulsivity. The indirect effect of treatment through change in craving was significant (self-reported use=-0.21, 95% Credible Interval [CrI]=-0.35, −0.09; drug screen-ascertained use=-0.36, 95% CrI=-0.63, −0.16). Change in craving mediated 56% of the treatment effect on self-reported use and 45% of the effect on drug screen-ascertained use. Estimates for mediated effects for depressive symptoms and impulsivity were smaller in magnitude and non-significant.

**Conclusion:** Reduction in craving mediates the effect of naltrexone-bupropion pharmacotherapy in methamphetamine use disorder. Craving may serve as a surrogate measure of treatment efficacy in short-term trials and help identify promising candidate medications to be tested in larger and longer-term trials.

**Trial Registration:** ClinicalTrials.gov number: NCT03078075.

## INTRODUCTION

The prevalence of methamphetamine use and methamphetamine use disorder (MUD) as well as fatal and nonfatal toxicity associated with such use in the United States has increased in the past two decades (Paulus and Stewart, 2020; Jones et al., 2022; Hoopsick et al., 2025; Chen et al., 2021; Han et al., 2021). For example, based on the United States Centers for Disease Control and Prevention, rates of methamphetamine-related deaths increased annually by 20.7% between 2008 and 2014 and by 32.2% between 2014 and 2021 (Hoopsick et al., 2025).

Over the years, numerous medications have been tested for treatment of MUD with mixed results. Some of these medications include non-selective serotonin reuptake inhibitor antidepressants, such as bupropion (Bakouni et al., 2023; Margolin et al., 1991; Newton et al., 2006; Ahmadi et al., 2019) and mirtazapine (Coffin et al., 2020; Naji et al., 2022), medications with stimulant properties, such as modafinil (Shearer et al., 2009; Elkrief et al., 2024; Anderson et al., 2012; Lee et al., 2013; Fard et al., 2020) and methylphenidate (Ling et al., 2014), as well as medications with efficacy for other substance use disorders such as buprenorphine and naltrexone (Ray et al., 2015; Ahmadi et al., 2019). However, there is currently no Food and Drug Administration (FDA) approved pharmacological treatment for MUD.

Notably, the potential biobehavioral mechanisms of action of these medications on MUD remain unclear. Shedding light on these mechanisms may help develop better and more individualized treatment approaches and determine whether prior candidate medications may have had selective benefits for key symptoms of MUD. Previous studies have identified meaningful reductions in craving, depressive symptoms, and impulsivity in the course of MUD treatment as potential mediators of significant treatment effect (Jha et al., 2025a; Jha et al., 2025b; Mojtabai et al., 2024; Ren et al., 2024; Liu et al., 2022; Hartz et al., 2001).

Craving has received growing attention in literature in recent years (Garbutt et al., 2016; Kleykamp et al., 2019; Garrison et al., 2023) and was added to the DSM-5 as a diagnostic criterion for substance use disorders partly based on the view that it “may become a biological treatment target” (Hasin et al., 2013). Craving has been closely linked with patterns of drug use in MUD treatment trials (Galloway et al., 2009; Huang et al., 2023; Kamp et al., 2019; Hartz et al., 2001; Mojtabai et al., 2024) and is sometimes used as a proxy indicator of treatment effect in short-term trials (Liu et al., 2022; Newton et al., 2006; Ahmadi et al., 2019).

Depression is also commonly associated with MUD and linked with poorer outcomes (Li et al., 2022; Leung et al., 2023; Glasner-Edwards et al., 2009). However, the association between change in depressive symptoms and drug use in past medication treatment trials for MUD has not been consistent (Naji et al., 2022; Elkashef et al., 2008; Coffin et al., 2020; Shoptaw et al., 2006).

Lastly, impulsivity, defined as “a tendency to act on the urge without forethought,”(Jha et al., 2025a) has been found to be relatively common among individuals with MUD and is considered a risk factor for poorer outcomes and return to drug use (Moallem et al., 2018; Hoffman et al., 2006; Mu et al., 2022; Luo et al., 2025).

These potential mechanisms were recently examined in secondary analyses of the Accelerated Development of Additive Pharmacotherapy Treatment for methamphetamine disorder (ADAPT-2) trial. ADAPT-2 was a two-stage randomized controlled trial (RCT) of extended-release injectable naltrexone (380 mg every 3 weeks) combined with oral bupropion (450 mg daily) (NTX-BUP) versus placebo (Trivedi et al., 2021). Using a two-stage design, placebo arm participants who did not achieve the study’s primary outcome were re-randomized to NTX-BUP versus placebo in the second Stage of the trial. The trial found a significant effect of NTX-BUP over placebo on the primary outcome, defined as at least 3 negative urine drug screens (UDS) out of four samples obtained at the end of Stage 1 or Stage 2.

In a set of secondary analyses limited to the first Stage of the ADAPT-2, lower levels of craving and impulsivity assessed each week in the course of the trial were found to be associated with higher probability of transitioning to negative urine toxicology at the next assessment point in the NTX-BUP group but not in the placebo group (Jha et al., 2025a). These analyses also found that higher levels of craving were associated with higher probability of a positive UDS at the next assessment in both the NTX-BUP and placebo groups. However, mediation was not examined in these analyses.

Another set of secondary analyses found that the slope of change in depressive symptoms in the first four weeks of the trial partially mediated the treatment effect on abstinence at the end of the trial (Jha et al., 2025b). However, these analyses were limited to the Stage 1 sample of ADAPT-2 and did not examine dynamic mediation during the course of the trial. Instead, indirect effect of treatment through one mediator variable (slope of change in depressive symptoms) on a single outcome (negative UDS at the end of trial) was examined.

In the present study, we build on this previous work (Jha et al., 2025a; Jha et al., 2025b) by conducting a comprehensive examination of the mediating effect of craving, depressive symptoms, and impulsivity, using data from Stages 1 and 2 of the ADAPT-2 trial. For these analyses, we used longitudinal multilevel structural equation modeling (LMSEM) which is specifically designed for intensive longitudinal data such as those produced in RCTs and captures dynamic effects of time-varying mediators and outcomes in the course of the trial (Berli et al., 2021).

This research can inform potential mechanisms underlying pharmacological treatment response in MUD. Furthermore, mediators that are closely linked with drug use patterns and are detectable early in the course of treatment may serve as proxy measures for screening and identifying promising candidate medications to be tested in larger and longer-term trials.

## METHODS

### Sampling and design

The sampling and design of ADAPT-2 have been previously described (Trivedi et al., 2021). Briefly, individuals aged 18-65 years with MUD who expressed interest in quitting or reducing methamphetamine use were recruited. To be eligible, participants had to meet the *Diagnostic and Statistical Manual of Mental Disorders*, fifth edition (DSM-5), criteria for moderate or severe MUD; report methamphetamine use on at least 18 of the 30 days prior to consent; have two or more methamphetamine-positive UDS within 10 days before randomization; and be free from opioids at randomization. Individuals receiving treatment for other substance use disorders, expected to need opioid medications, using medications contraindicated with study medications, or with conditions increasing the risk of seizure were excluded (Trivedi et al., 2021).

The trial used a two-stage, sequential parallel comparison design (SPCD) (Fava et al., 2003). In Stage 1, participants were assigned based on a 0.26:0.74 ratio to the NTX-BUP or matched injectable plus oral placebo arm. In Stage 2, participants in the placebo arm who did not achieve the study’s primary outcome (i.e., placebo non-responders) were re-randomized based on a 1:1 ratio to active treatment or placebo. The purpose of this design is to maximize the power of the study compared to a one-stage parallel comparison RCT and to exclude placebo responders from the Stage 2 sample (Fava et al., 2003; Fava, 2024; Song et al., 2025). As described by Fava, “Stage 1 of SPCD is aimed at comparing drug and placebo in a standard parallel comparison design fashion, with drug-placebo differences being expected to be standard, while generating a large cohort of placebo non-responders for the second stage (this is why the randomization ratio favors placebo). Stage 2 is aimed at comparing drug and placebo only among Stage 1 placebo nonresponders, with drug-placebo differences being expected to be greater and placebo responses being markedly lower, as these patients on Stage 2 have already ‘failed placebo’.” (p. 200, Fava, 2024) The trial was conducted in accordance with the principles of the Declaration of Helsinki.

The study protocol was approved by the Data and Safety Monitoring Board of the National Institute on Drug Abuse (NIDA) Clinical Trials Network, by a central institutional review board, and by institutional review boards at study sites (ClinicalTrials.gov number, NCT03078075) (Trivedi et al., 2021). Informed consent was obtained from all participants. This secondary analysis used deidentified and publicly available data deemed exempt from review by the institutional review board of Tulane University. Data are publicly available from the NIDA datashare site (https://datashare.nida.nih.gov/study/nida-ctn-0068, principal investigator: Madhukar Trivedi, MD).

### Assessments

*Urine drug screen (UDS)* for qualitative methamphetamine results was collected twice a week. The integrity of urine samples was determined by using an embedded temperature strip on the collection cup and testing for adulterants.

*Self-reported use* of methamphetamine was assessed using the Timeline Followback (TLFB) (Sobell, 1992; Fals-Stewart et al., 2000). TLFB was administered twice a week. At each administration, TLFB assessed drug use on each day since the previous TLFB administration.

*Craving* was assessed weekly using a visual analogue scale to capture the most severe methamphetamine craving during the previous week, with values ranging from 0 to 100 (McHugh et al., 2014; Lee et al., 2002). Higher values indicate higher levels of craving.

*Depressive symptoms* were assessed weekly using the nine-item Patient Health Questionnaire (PHQ-9) (Kroenke et al., 2001). Participants rated the frequency of symptoms on a 4-point Likert scale from 0 (“never”) to 3 (“nearly every day”). Total PHQ-9 scores range from 0 to 27. Higher values indicate higher levels of depressive symptoms.

*Impulsivity* was assessed using the sum of scores on two items on the impulsivity subscale of the *Concise Health Risk Tracking* (CHRT) scale, administered weekly (Trombello et al., 2023). The items ask about “saying or doing things without thinking” and the frequency of making decisions “quickly or ‘on impulse’” in the past week. Responses are on a Likert scale ranging from 0 (“strongly disagree”) to 4 (“strongly agree”). Total score on the scale ranges from 0 to 8. Higher values indicate greater levels of impulsivity. Construct validity and reliability of this measure have been examined in past research (Trombello et al., 2023).

Additionally, models adjusted for age, sex, race-ethnicity, other substance use disorders, and alcohol use disorder. Race/ethnicity was dichotomized to non-Hispanic white versus minority groups for these analyses. Alcohol and other substance (opioid, cocaine, cannabis and sedative) use disorders (any versus none) were ascertained at baseline based on DSM-5 criteria.

### Analytic Approach

In preliminary analyses, we examined time trends in outcomes and potential mediators using a series of logistic (for binary variables) and linear (for continuous variables) general estimation equation (GEE) models clustered within participant, with the interaction of time (in weeks) and treatment arm included in the models. Marginal effects were computed and graphed.

Next, we used LMSEM to examine dynamic mediation of treatment effect by craving, depressive symptoms, and impulsivity separately for the self-reported and UDS-ascertained methamphetamine use. These LMSEM models were based on the model adapted by Berli and colleagues (Berli et al., 2021) who extended the MSEM model originally described by Preacher and colleagues (Preacher et al., 2010), by adding temporal effects. This LMSEM model allows for explicit representation of direct and indirect paths and for within-person processes that vary across participants.

Similar to Berli and colleagues (Berli et al., 2021), we fitted the LMSEM model using a two-level random effects specification. We allowed for random effects for time (days since randomization) on both the mediators and the outcomes and the within-person association of time-varying mediators with the outcomes. The indirect effect was calculated by multiplying the regression coefficient for mediator on treatment arm by the combined average within-person and contextual effects of the outcome on the mediator (Pituch and Stapleton, 2012).

As in Berli and colleagues’ study (Berli et al., 2021), in the main analyses, we did not lag the mediator variables because craving, depressive symptoms, and impulsivity were assessed for the preceding week and were thus inherently lagged. Nonetheless, in sensitivity analyses, we additionally examined one-day lagged effects of these mediators.

As both outcomes (self-reported and UDS-ascertained methamphetamine use) are binary, they were modeled with a probit link, which allows for computing the total effect as the summation of the direct and indirect effects (MacKinnon and Dwyer, 1993).

*Mplus* 8.11 with Bayesian estimator was used for the mediation analyses (see Appendix A for more detail) (Geiser, 2021). Six models were run in total—one for each combination of the three mediators and the two outcomes. Demographic characteristics, alcohol and other substance use disorder comorbidity, and the intervention arm were entered into each model as time invariant covariates. Similar to the main analyses of the trial (Trivedi et al., 2021), data from both Stages were included. In addition, a dummy variable for Stage was entered into the models to account for differences in the average values of the mediators and the probabilities of the outcomes across Stages.

The ADAPT-2 data include two levels of analytic dependence: 1) clustering of assessments from the same individuals over time, and 2) inclusion of some participants from Stage 1 in Stage 2. *Mplus*, however, does not allow for more than 1 level of clustering. In a set of secondary analyses we adjusted the standard errors of the models for overlap of participants from Stage 1 and Stage 2 using a design-effect adjustment factor (Kish, 1965). Separate adjustment factors were computed for self-reported drug use and UDS-ascertained drug use. New credible intervals were computed in sensitivity analysis based on these adjustment factors. Similar to the full maximum likelihood estimation method, Bayesian estimation in *Mplus* uses all available information (Asparouhov, 2010). As such, it is robust to the effect of data missing at random. Fully 90.0% of participants of the trial were maintained in the study for 6 weeks or longer. Those who dropped out earlier were evenly distributed across the study weeks (Appendix B).

## RESULTS

### Trial Outcomes

A total of 403 participants were randomized in Stage 1, including 109 (27.0%) assigned to NTX-BUP and 294 (73.0%) to placebo; 225 were re-randomized in Stage 2, including 114 (50.7%) assigned to NTX-BUP and 111 (49.3%) to placebo. Thus, the effective sample was 223 for NTX-BUP and 405 for placebo.

Socio-demographic characteristics of the participants were previously reported in detail (Trivedi et al., 2021). Briefly, the mean age of participants was 41 (standard deviation [SD]=10.1) years, 68.7% were male, and 67.7% were Non-Hispanic white. On average, participants used methamphetamine on 27 of the 30 days before the consent day (Trivedi et al., 2021).

Consistent with other primary and secondary analyses based on this trial (Jha et al., 2025a; Jha et al., 2025b; Trivedi et al., 2021), we replicated the significant effects of the treatment arm and time trends for all outcomes and potential mediators in GEE analyses (Appendices C-G). Additionally, the coefficients for interaction of treatment arm and time were statistically significant in the models for UDS-ascertained drug use (Appendix D), craving (Appendix E), and depressive symptoms (Appendix F).

### Mediation Analyses

In LMSEM analyses, all models with craving and depressive symptoms as mediators, as well as the model for self-reported drug use with impulsive behavior as a mediator, converged normally. However, the model for UDS-ascertained methamphetamine use with impulsivity as a mediator only converged after it was simplified by specifying fixed slopes for time.

In these analyses, the indirect effects of treatment on outcomes via craving were significant (estimate=-0.21, 95% Credible Interval [CrI]= −0.35, −0.09, for self-reported methamphetamine use; estimate= −0.36, 95% CrI= −0.63, −0.16, for UDS-ascertained methamphetamine use) (Table 1, Figure 1A and 1B). 56% of the effect of treatment on self-reported use and 45% of the effect of treatment on UDS-ascertained use were mediated by change in craving.

**Table 1:** Parameter estimates from the longitudinal multilevel structural equation mediation models testing the mediating effect of craving in Accelerated Development of Additive Pharmacotherapy Treatment for methamphetamine disorder (ADAPT-2) randomized controlled trial of naltrexone-bupropion (NTX-BUP) treatment according to the type of outcome.

| Effects | Self-reported methamphetamine use |  | Urine drug screen-ascertained methamphetamine use |  |
| --- | --- | --- | --- | --- |
|  | Estimate | 95% CrI<sup>a</sup> | Estimate | 95% CrI<sup>a</sup> |
| **Within-person effects<sup>b</sup>** |  |  |  |  |
| Effect of time on methamphetamine use | 0.04 | 0.01, 0.06<sup>c</sup> | 0.07 | -0.01, 0.15 |
| Effect of time on craving | -0.23 | -0.25, -0.20<sup>c</sup> | -0.23 | -0.25, -0.20 |
| Effect of craving on methamphetamine use | 0.21 | 0.14, 0.28<sup>c</sup> | 0.40 | 0.31, 0.48<sup>c</sup> |
| **Between-person effects<sup>b</sup>** |  |  |  |  |
| Effect on methamphetamine use of... |  |  |  |  |
| Treatment arm (NTX-BUP versus placebo) | -0.16 | -0.42, 0.09 | -0.45 | -0.88, 0.01 |
| Craving | 0.01 | 0.01, 0.02<sup>c</sup> | 0.02 | 0.01, 0.03<sup>c</sup> |
| Female | 0.14 | -0.12, 0.39 | 0.80 | 0.33, 1.31<sup>c</sup> |
| Age in years | 0.02 | 0.01, 0.03<sup>c</sup> | 0.01 | -0.01, 0.03 |
| Minority race-ethnicity | 0.26 | 0.00, 0.52<sup>c</sup> | 0.26 | -0.21, 0.73 |
| Other substance use disorder<sup>d</sup> | -0.10 | -0.35, 0.14 | 0.17 | -0.29, 0.64 |
| Alcohol use disorder<sup>e</sup> | -0.15 | -0.44, 0.12 | -0.49 | -1.02, 0.04 |
| Trial Stage | -0.23 | -0.48, 0.02 | 0.10 | -0.38, 0.57 |
| Effect on craving of... |  |  |  |  |
| Treatment arm (NTX-BUP versus placebo) | -6.68 | -10.60, -2.91<sup>c</sup> | -6.88 | -10.75, -3.15<sup>c</sup> |
| Female sex | -1.25 | -5.17, 2.80 | -1.25 | -5.25, 2.70 |
| Age in years | -0.21 | -0.40, -0.04<sup>c</sup> | -0.22 | -0.40, -0.05<sup>c</sup> |
| Minority race-ethnicity | -5.58 | -9.55, -1.71<sup>c</sup> | -5.87 | -9.64, -1.89<sup>c</sup> |
| Other substance use disorder<sup>d</sup> | 0.65 | -3.23, 4.58 | 0.90 | -2.89, 4.75 |
| Alcohol use disorder<sup>e</sup> | -1.26 | -5.72, 3.18 | -1.42 | -5.98, 3.17 |
| Trial Stage | -12.04 | -15.86, -8.07<sup>c</sup> | -11.88 | -15.82, -7.97<sup>c</sup> |
| Indirect effect of treatment (mediated)<sup>f</sup> | -0.21 | -0.35, -0.09<sup>c</sup> | -0.36 | -0.63, -0.16<sup>c</sup> |
| Ratio of indirect effect over total effect of treatment (direct+indirect) | 0.56 | -- | 0.45 | -- |

<sup>a</sup>.CrI stands for credible interval.
<sup>b</sup>.The within-person results are based on standardized estimates averaged over persons; between-person results are based on unstandardized estimates.
<sup>c</sup>.Significant estimate (i.e., credible interval does not include 0).
<sup>d</sup>.Based on DSM-5 checklist administered at baseline (includes opioids, cocaine, cannabis and sedatives).
<sup>e</sup>.Based on DSM-5 checklist administered at baseline.
<sup>f</sup>. Computed as the effect of mediator on treatment multiplied by the sum of within-person and contextual effects of mediator on methamphetamine use.

**Figure 1A:**
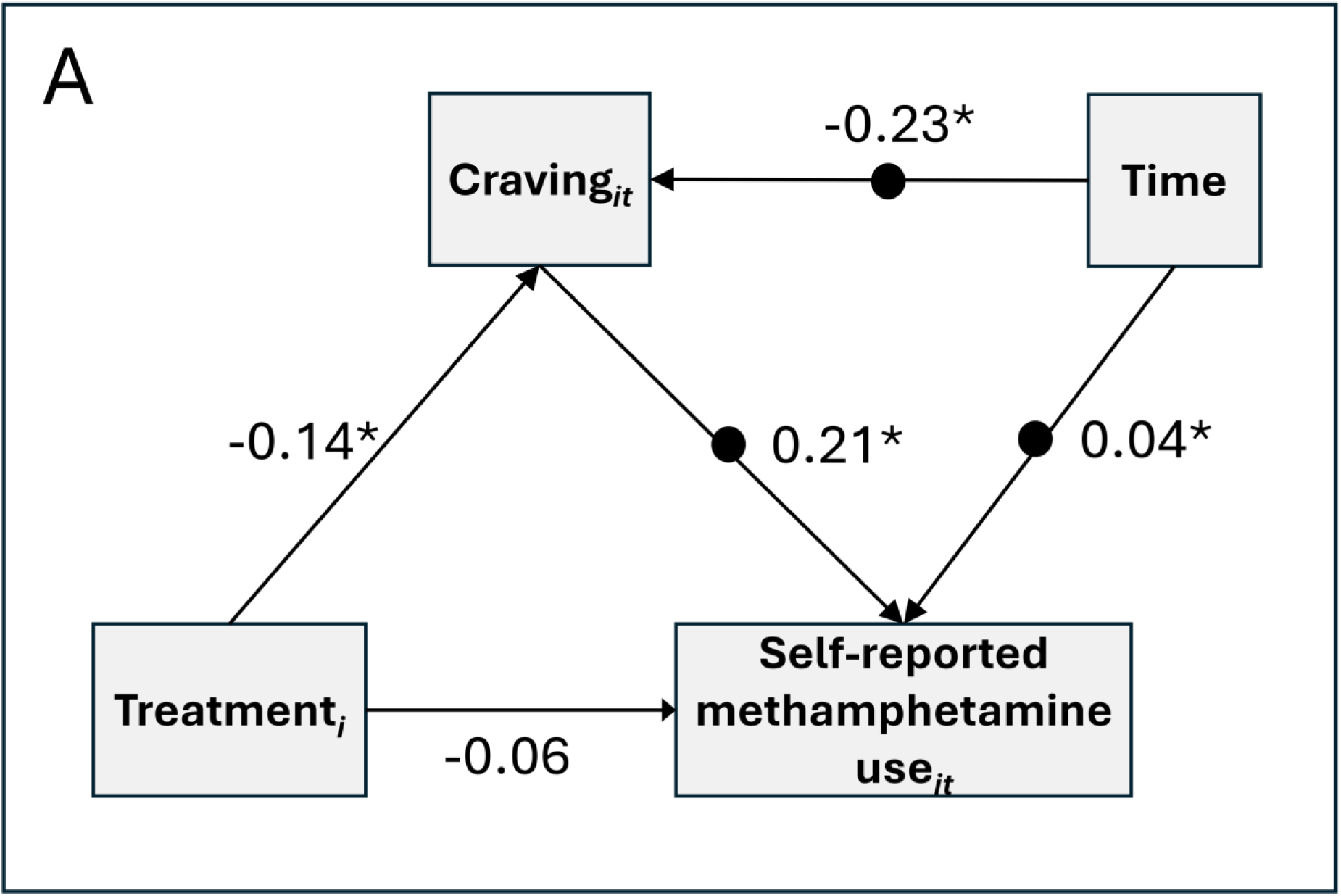
Longitudinal multilevel structural equation mediation model of naltrexone-bupropion (NTX-BUP) treatment effect in Accelerated Development of Additive Pharmacotherapy Treatment for methamphetamine disorder (ADAPT-2) randomized controlled trial on self-reported use of methamphetamine as mediated by craving. Random within-person effects of craving and time are presented as black circles on the paths. Presented results are based on within-person standardized estimates averaged over persons. Subscript *i* stands for individual and *t* for time.

**Figure 1B:**
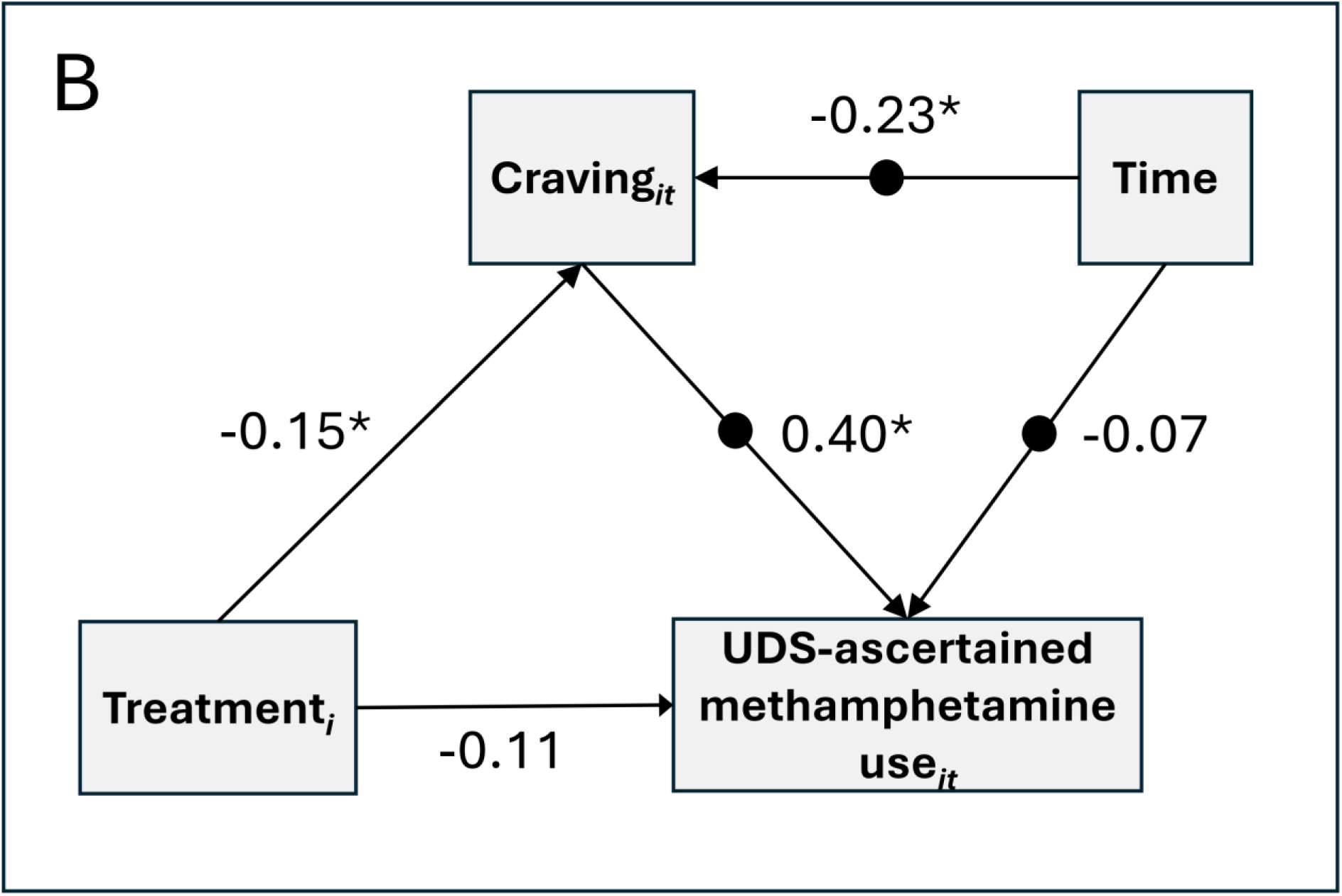
Longitudinal multilevel structural equation mediation model of naltrexone-bupropion (NTX-BUP) treatment effect in Accelerated Development of Additive Pharmacotherapy Treatment for methamphetamine disorder (ADAPT-2) randomized controlled trial on urine drug screen (UDS) ascertained use of methamphetamine as mediated by craving. Random within-person effects of craving and time are presented as black circles on the paths. Presented results are based on within-person standardized estimates averaged over persons. Subscript *i* stands for individual and *t* for time.

The indirect effects via depressive symptoms were smaller in magnitude and non-significant (estimate= −0.02, 95% CrI= −0.07, 0.01, for self-reported use; estimate= −0.06, 95% CrI= −0.19, 0.03, for UDS-ascertained use). Similarly, the indirect effects via impulsivity were relatively small and non-significant (estimate=,-0.04, 95% CrI=,-0.11, 0.03, for self-reported use; estimate= −0.09, 95% CrI= −0.26, 0.07 for UDS-ascertained use) (Tables 2 and 3).

**Table 2:** Parameter estimates from the longitudinal multilevel structural equation mediation models testing the mediating effect of depressive symptoms in Accelerated Development of Additive Pharmacotherapy Treatment for methamphetamine disorder (ADAPT-2) randomized controlled trial of naltrexone-bupropion (NTX-BUP) treatment according to the type of outcome.

| Effects | Self-reported methamphetamine use |  | Urine drug screen-ascertained methamphetamine use |  |
| --- | --- | --- | --- | --- |
|  | Estimate | 95% CrI<sup>a</sup> | Estimate | 95% CrI<sup>a</sup> |
| **Within-person effects<sup>b</sup>** |  |  |  |  |
| Effect of time on methamphetamine use | -0.01 | -0.04, 0.01 | -0.01 | -0.01, 0.08 |
| Effect of time on depressive symptoms | -0.13 | -0.16, -0.11<sup>c</sup> | -0.13 | -0.16, -0.10<sup>c</sup> |
| Effect of depressive symptoms on methamphetamine use | 0.05 | -0.02, 0.12 | 0.23 | 0.14, 0.31<sup>c</sup> |
| **Between-person effects<sup>b</sup>** |  |  |  |  |
| Effect on methamphetamine use of... |  |  |  |  |
| Treatment arm (NTX-BUP versus placebo) | -0.22 | -0.45, 0.01 | -0.50 | -0.88, -0.11<sup>c</sup> |
| Craving | 0.02 | -0.00, 0.04 | 0.03 | -0.01, 0.06 |
| Female | 0.10 | -0.14, 0.34 | 0.67 | 0.25, 1.11<sup>c</sup> |
| Age in years | 0.01 | 0.00, 0.03<sup>c</sup> | 0.01 | -0.01, 0.03 |
| Minority race-ethnicity | 0.15 | -0.09, 0.37 | 0.11 | -0.28, 0.50 |
| Other substance use disorder<sup>d</sup> | -0.11 | -0.33, 0.12 | 0.15 | -0.26, 0.56 |
| Alcohol use disorder<sup>e</sup> | -0.15 | -0.42, 0.11 | -0.41 | -0.87, 0.04 |
| Trial Stage | -0.35 | -0.59, -0.13<sup>c</sup> | -0.07 | -0.46, 0.32 |
| Effect on depressive symptoms of... |  |  |  |  |
| Treatment arm (NTX-BUP versus placebo) | -0.64 | -1.58, 0.32 | -0.64 | -1.56, 0.29 |
| Female sex | -0.30 | -1.25, 0.68 | -0.28 | -1.23, 0.69 |
| Age in years | -0.07 | -0.11, -0.03<sup>c</sup> | -0.07 | -0.11, -0.03<sup>c</sup> |
| Minority race-ethnicity | 0.15 | -0.81, 1.09 | 0.12 | -0.83, 1.07 |
| Other substance use disorder<sup>d</sup> | 1.71 | 0.77, 2.64<sup>c</sup> | 1.71 | 0.80, 2.65<sup>c</sup> |
| Alcohol use disorder<sup>e</sup> | 0.02 | -1.07, 1.07 | -0.01 | -1.08, 1.06 |
| Trial Stage | -1.01 | -1.93, -0.07<sup>c</sup> | -1.03 | -1.94, 0.08<sup>c</sup> |
| Indirect effect<sup>f</sup> | -0.02 | -0.07, 0.01 | -0.06 | -0.19, 0.03 |
| Ratio of indirect effect over total effect of treatment (direct+indirect) | 0.09 | -- | 0.12 | -- |

<sup>a</sup>. CrI stands for credible interval.
<sup>b</sup>. The within-person results are based on standardized estimates averaged over persons; between-person results are based on unstandardized estimates.
<sup>c</sup>. Significant estimate (i.e., credible interval does not include 0).
<sup>d</sup>. Based on DSM-5 checklist administered at baseline (includes opioids, cocaine, cannabis and sedatives).
<sup>e</sup>. Based on DSM-5 checklist administered at baseline.
<sup>f</sup>. Computed as the effect of mediator on treatment multiplied by the sum of within-person and contextual effects of mediator on methamphetamine use.

**Table 3:** Parameter estimates from the longitudinal multilevel structural equation mediation models testing the mediating effect of impulsivity in Accelerated Development of Additive Pharmacotherapy Treatment for methamphetamine disorder (ADAPT-2) randomized controlled trial of naltrexone-bupropion (NTX-BUP) treatment according to the type of outcome.

| Effects | Self-reported methamphetamine use |  | Urine drug screen-ascertained methamphetamine use |  |
| --- | --- | --- | --- | --- |
|  | Estimate | 95% CrI<sup>a</sup> | Estimate | 95% CrI<sup>a</sup> |
| **Within-person effects<sup>b</sup>** |  |  |  |  |
| Effect of time on methamphetamine use | -0.00 | -0.03, 0.02 | -0.00 | -0.08, 0.01 |
| Effect of time on impulsivity | -0.13 | -0.15, -0.10<sup>c</sup> | -0.13 | -0.15, -0.10<sup>c</sup> |
| Effect of impulsivity on methamphetamine use | 0.10 | 0.04, 0.17<sup>c</sup> | 0.27 | 0.18, 0.34<sup>c</sup> |
| **Between-person effects<sup>b</sup>** |  |  |  |  |
| Effect on methamphetamine use of... |  |  |  |  |
| Treatment arm (NTX-BUP versus placebo) | -0.22 | -0.46, 0.02 | -0.48 | -0.87, -0.11<sup>b</sup> |
| Craving | 0.08 | 0.03, 0.14<sup>c</sup> | 0.16 | 0.07, 0.26<sup>b</sup> |
| Female | 0.10 | -0.14, 0.34 | 0.68 | 0.26, 1.13<sup>b</sup> |
| Age in years | 0.02 | 0.00, 0.03<sup>c</sup> | 0.01 | -0.01, 0.03 |
| Minority race-ethnicity | 0.18 | -0.05, 0.41 | 0.16 | -0.24, 0.56 |
| Other substance use disorder<sup>d</sup> | -0.14 | -0.37, 0.09 | 0.08 | -0.32, 0.49 |
| Alcohol use disorder<sup>e</sup> | -0.16 | -0.42, 0.11 | -0.41 | -0.87, 0.05 |
| Trial Stage | -0.34 | -0.58, -0.11<sup>c</sup> | -0.05 | -0.44, 0.34 |
| Effect on impulsivity of... |  |  |  |  |
| Treatment arm (NTX-BUP versus placebo) | -0.21 | -0.58, 0.15 | -0.20 | -0.57, 0.16 |
| Female sex | 0.02 | -0.36, 0.40 | 0.02 | -0.34, 0.42 |
| Age in years | -0.02 | -0.04, -0.01<sup>c</sup> | -0.02 | -0.04, -0.01<sup>c</sup> |
| Minority race-ethnicity | -0.35 | -0.73, 0.02 | -0.36 | -0.73, 0.00 |
| Other substance use disorder<sup>d</sup> | 0.72 | 0.35, 1.08<sup>c</sup> | 0.73 | 0.36, 1.09<sup>c</sup> |
| Alcohol use disorder<sup>e</sup> | -0.00 | -0.42, 0.41 | -0.02 | -0.46, 0.41 |
| Trial Stage | -0.54 | -0.90, -0.17<sup>c</sup> | -0.54 | -0.91, -0.19<sup>c</sup> |
| Indirect effect<sup>f</sup> | -0.04 | -0.11, 0.03 | -0.09 | -0.26, 0.07 |
| Ratio of indirect effect over total effect of treatment (direct+indirect) | 0.13 | -- | 0.15 | -- |

<sup>a</sup>. CrI stands for credible interval.
<sup>b</sup>. The within-person results are based on standardized estimates averaged over persons; between-person results are based on unstandardized estimates.
<sup>c</sup>. Significant estimate (i.e., credible interval does not include 0).
<sup>d</sup>. Based on DSM-5 checklist administered at baseline (includes opioids, cocaine, cannabis and sedatives).
<sup>e</sup>. Based on DSM-5 checklist administered at baseline.
<sup>f</sup>. Computed as the effect of mediator on treatment multiplied by the sum of within-person and contextual effects of mediator on methamphetamine use.

Because an earlier analyses of ADAPT-2 (Jha et al., 2025b) had identified a significant mediation effect for depressive symptoms, we conducted further analyses to explore whether the discrepant results between our study and the previous study could be attributed to differences in analytic methods or differences in samples used. The mediation analysis in the previous study was based on the slope of change in depressive symptoms in the first four weeks of the trial, examined outcome at the end of trial and was limited to the first Stage of the trial.

Therefore, in these further analyses, we limited the sample to the first Stage of ADAPT-2 trial and used LMSEM without random effects for time. The indirect effect via depressive symptoms on UDS-ascertained methamphetamine use in this analysis was significant (estimate= −0.13, 95% CrI= −0.30, −0.02), although it was much smaller than the direct effect (estimate = −0.76, 95% CrI= −1.23, −0.30), accounting for approximately 15% of the total effect. In comparison, 24.8% of the total effect was estimated to be mediated by depressive symptoms in Jha and colleagues’ study (Jha et al., 2025b).

### Sensitivity Analyses

The results of the sensitivity analysis for lagged effects of craving as a mediator (indirect effects) were similar to the main analysis for self-reported methamphetamine use (estimate= −0.17, 95% CrI= −0.31, −0.07; 53% of the effect mediated) and UDS-ascertained use (estimate= −1.28, 95% CrI= −2.34, −0.46, 54% of the effect mediated). To achieve convergence, a fixed slope had to be used for the effect of time in the lagged model for UDS-ascertained methamphetamine use. The lagged model for the indirect effect of depressive symptoms on self-reported methamphetamine use did not find a significant result and the model for UDS-ascertained methamphetamine use did not converge. The lagged models for impulsivity similarly did not find a significant mediating effect on self-reported or UDS-ascertained methamphetamine use (data not shown).

The results of sensitivity analysis adjusting for overlap in participants from both Stages of ADAPT-2 were substantively concordant with the results of the main analyses. Craving continued to be a significant mediator of the treatment effect for both self-reported methamphetamine use (estimate= −0.31, 95% CrI= −0.57, −0.06, 61% of the effect mediated) and for UDS-ascertained methamphetamine use (estimate = −0.51, 95% CrI= −0.97, −0.05, 54% of the effect mediated). The estimates of the indirect effects in these models were somewhat different than the estimates in the main analyses because the fixed effect for Stage was not included in these models. The indirect effects for depressive symptoms and impulsivity were non-significant in these sensitivity analyses (data not shown).

## DISCUSSION

Understanding the biobehavioral mechanisms underlying the treatment effect is essential for drug development in neuropsychiatry. Research on mechanisms of treatment effect also has implications for identifying factors that contribute to return to use and recovery from substance use disorders. However, there is scant research on treatment mechanisms underlying potential pharmacotherapies for stimulant use disorders. In the present secondary analyses of the ADAPT-2 trial, we used the LMSEM method to examine longitudinal mediating effects of craving, depressive symptoms and impulsivity in an RCT that had identified a positive signal for NTX-BUP as a potential treatment of MUD. Our analyses found that craving—but not depressive symptoms or impulsivity—significantly mediated the effect of NTX-BUP treatment for MUD as determined through both self-reported and UDS-ascertained methamphetamine use.

Among these three potential mediators, only the effect of depressive symptoms had previously been investigated (Jha et al., 2025b). The Jha and colleagues’ study, however, differed from our analyses in several ways: it was limited to Stage 1 of ADAPT-2 trial, it defined the mediator as change in depressive symptoms from baseline to week 4 and the outcome as negative results in 3 out of 4 UDS for methamphetamine in weeks 5 and 6 of the trial, and the outcome was limited to UDS-ascertained use (Jha et al., 2025b). This design is unlikely to capture the effect of fluctuations in mood in short spans of time which have been shown to be related to drug use patterns in stimulant use disorders (Montoya et al., 2013).

In contrast, we used the dynamic LMSEM in which the effect of weekly fluctuations in mediators on the weekly measured outcomes were examined. Furthermore, similar to the main ADAPT-2 study report, we used data from both Stages of the trial. As such, the results of the mediation analysis are more closely tied to the results of the main ADAPT-2 trial. Probably because of these differences in sampling and analytic approach, the results of our study were different than the results of Jha and colleauges’ study (Jha et al., 2025b) which found a significant mediating effect for depressive symptoms.

To ascertain whether differences across the two studies could be attributed to differences in the samples, we repeated our analyses after limiting the sample to the first Stage of ADAPT-2. In these repeat analyses we did find a significant, albeit a smaller mediation effect than the one found in Jha and colleagues’ study, suggesting the mediating effect of depressive symptoms may be more prominent earlier in the course of the trial (i.e., Stage 1). This finding is puzzling because the effect of treatment on the primary study outcome and on depressive symptoms, as well as the baseline PHQ-9 scores, were similar in the two Stages of ADAPT-2 trial (Trivedi et al., 2021). In view of the high prevalence of depressive symptoms in individuals with MUD and the association of these symptoms with poorer outcomes, further research on the potential mediating role of these symptoms is warranted.

Our study is also the first to examine the mediating effect of craving and impulsivity in the ADAPT-2 trial, building off a prior secondary analysis of ADAPT-2 that examined the effect of craving and impulsivity on UDS-ascertained methamphetamine use and the moderating effect of NTX-BUP treatment on these associations in the Stage 1 of ADAPT-2 trial (Jha et al., 2025a). However, no mediation analysis were conducted in that study. As such, the results of Jha and colleagues’ study cannot be compared with our study results.

Importantly, the findings of our study extend past research on the role of craving in persistence of methamphetamine use in individual with MUD (Galloway et al., 2009; Huang et al., 2023; Kamp et al., 2019; Hartz et al., 2001; Mojtabai et al., 2024) and the effect of naltrexone on cue-induced craving in MUD (Roche et al., 2017). For example, in a prospective, repeated-measures study of adults with MUD, weekly ratings of craving intensity significantly predicted methamphetamine use in the subsequent week (Hartz et al., 2001). Craving scores in the week preceding methamphetamine use in this study were 2.7 times higher than those preceding non-use (Hartz et al., 2001). The authors concluded that craving is a salient predictive factor in continued methamphetamine use in patients receiving treatment for MUD. The present study extends this conclusion by suggesting that effective pharmacological treatment of MUD may work, at least in part, by attenuating craving levels.

The findings should be considered in light of several limitations. First, because of the relatively small number of assessments of UDS and mediators, we could not use dynamic structural equation modeling (DSEM) which accounts for autoregressive associations (Hamaker et al., 2018). Similarly, because of these data limitations, we were unable to examine the interaction of treatment with time or the potential impact of change in methamphetamine use on subsequent craving, depressive symptoms, and/or impulsivity. Reciprocal relationships between stimulant drug use and craving are reported in both pre-clinical and clinical studies (Shelton and Beardsley, 2008; Mahoney et al., 2007) and should be examined in future research. Second, past research has found significant correlations between craving, depressive symptoms, and impulsivity among individuals with MUD (Tziortzis et al., 2011; Shen et al., 2012; Nakama et al., 2008), suggesting possible overlap in their effects. However, the LMSEM models could not accommodate multiple mediators simultaneously. Third, the ADAPT-2 trial was not designed for mediation analysis, and these secondary analyses might have been underpowered to detect smaller effects, therefore future replication efforts are warranted. Fourth, investigators have noted several limitations of SPCD model for estimating population effect of treatments (Song et al., 2025; Fava, 2024). Importantly, the target population for these RCTs is comprised of intention-to-treat subjects in Stage 1 and placebo non-responders in Stage 2. These populations may differ with regard to baseline variables and certainly differ in their responses to treatment.

Therefore, the results of these RCTs may not generalize to the general population of persons seeking treatment. Another limitation is the possibility of misclassification of placebo non-responders when using somewhat arbitrary categorical criteria to identify non-response (such as 3 negative UDS results at the end of trial). While these limitations would impact generalizability and interpretation of efficacy results from studies using SPCD, they may be less critical for our analyses which focused on the mechanisms of treatment response, and not generalizability of treatment efficacy. Furthermore, we assessed a dynamic outcome during the course of the trial rather than using an arbitrary cutoff to define outcome. Fifth, a limited number of outcome variables were measured weekly in ADAPT-2 trial, thus limiting the range of treatment outcomes we could assess. Future research needs to examine the mediators of treatment effect for other outcomes, especially quality of life and social integration. However, changes in these outcomes may not be detectable in short pharmacotherapy trials and may require longer treatment.

In conclusion, the present findings highlight the potential role of reduction in craving as a primary biobehavioral mechanism of action of NTX-BUP treatment for MUD. The findings also highlight the value of reduction in craving as a potential surrogate measure for assessing treatment effect in MUD trials. Craving was added to the DSM-5 as a diagnostic criterion for substance use disorders partly based on its promise as a biological treatment target (Hasin et al., 2013). Evidence from this study supports this notion. Ultimately, craving may serve as a treatment target in trials that are not long enough or have sufficient power to detect abstinence or prolonged changes in drug use patterns. Efficacy signals detected through this surrogate measure may elucidate promising candidate medications to be tested in more definitive trials.

## STATEMENTS AND DECLARATIONS

### Ethical Considerationsh

The Ethics Committee of Tulane University waived the need for ethics approval and patient consent for analysis and publication of publicly available anonymised data for this study (not human research).

### Consent to Participate

The Ethics Committee of Tulane University waived the need for ethics approval and patient consent for analysis and publication of publicly available anonymised data for this study (not human research).

### Consent for Publication

Not applicable.

### Author Contributions

Dr. Mojtabai had full access to all of the data in the study and takes responsibility for the integrity of the data and the accuracy of the data analysis.

*Concept and design:* Mojtabai, Amin-Esmaeili, Susukida.

*Acquisition, analysis, or interpretation of data*: Mojtabai, Amin-Esmaeili, Susukida, Nguyen.

*Drafting of the manuscript:* All authors.

*Critical review of the manuscript for important intellectual content*: All authors.

*Statistical analysis*: Mojtabai, Amin-Esmaeili, Susukida, Nguyen.

*Obtained funding:* Mojtabai, Amin-Esmaeili, Susukida, Nguyen, Dunn.

*Administrative, technical, or material support:* Mojtabai, Amin-Esmaeili, Susukida.

### Data Availability

This study is based on secondary analysis of publicly available data. All data files and codebooks can be downloaded from https://datashare.nida.nih.gov/study/nida-ctn-0068.

## Appendices

## Appendix A Description of Bayesian Estimation

Bayesian estimators were computed with two Markov Chain Monte Carlo (MCMC) chains with a maximum of 25,000 iterations in each chain. As most models converged in less than 3,000 iterations, we set the minimum number of iterations at 6,000, with the first 3,000 iterations as “burn-in”. A thinning factor of 30 was applied such that every 30th iteration would be used in the computation of the final posterior parameter distributions.

Convergence of the models was assessed by examining Bayesian trace plots and Potential Scale Reduction (PSR) values. PSR is based on a comparison of the parameter variation within each chain and across multiple iterations to the parameter variation across chains. Trace plots forming a relatively tight horizontal band with no large discrepancies between the two chains, at least in the post “burn-in” phase, and a stable PSR<1.05 indicate good convergence. Further iterations after reaching this PSR value are to ensure PSR does not increase in later iterations (Geiser, 2021).

## Appendix B

Distribution of participants in Additive Pharmacotherapy Treatment for methamphetamine disorder (ADAPT-2) randomized controlled trial according to the number of weeks they remained in the trial (average across Stages).

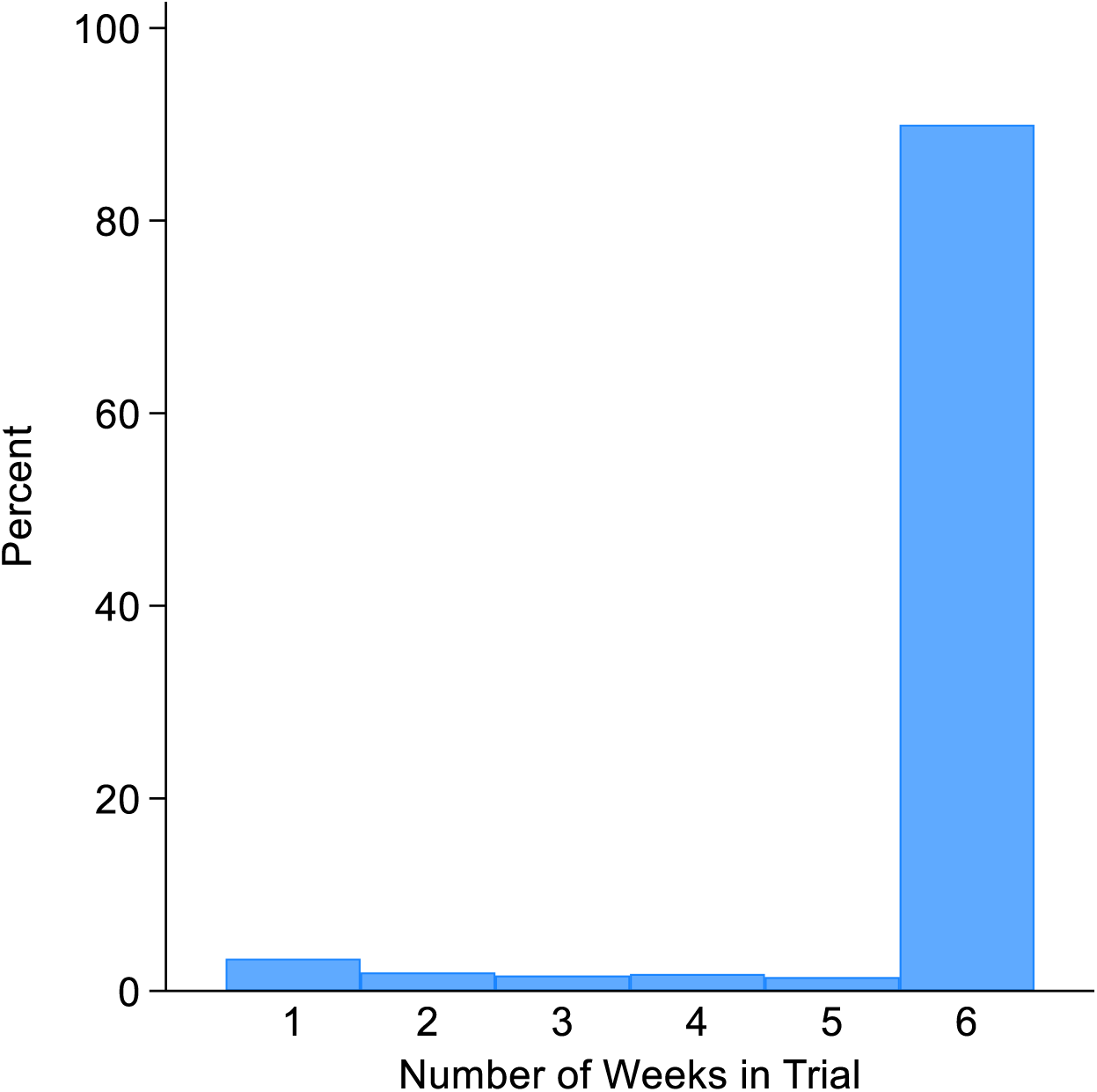

## Appendix C

Predicted average weekly probability of self-reported methamphetamine use in Additive Pharmacotherapy Treatment for methamphetamine disorder (ADAPT-2) randomized controlled trial of naltrexone-bupropion (NTX-BUP) versus placebo.

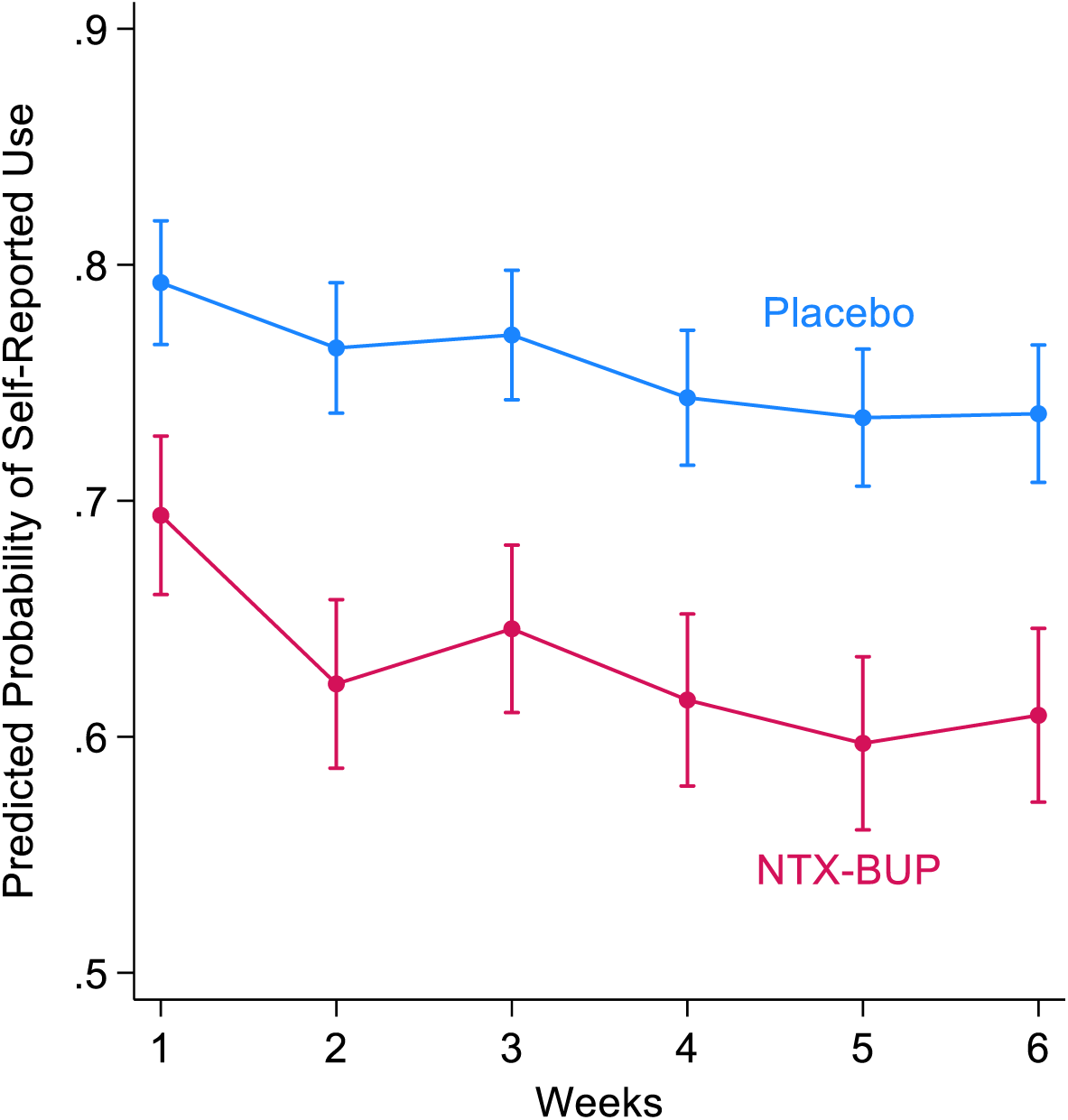

| Generalized estimation equation analysis results for the association of treatment arm, time and treatment arm by time interaction with self-reported use of methamphetamine in the course trial (logit link). |  |  |  |
| --- | --- | --- | --- |
| Variable | Regression Coefficient | 95% Confidence Interval | *p* |
| NTX-BUP arm (versus placebo) | -0.59 | -0.72, -0.45 | <0.001 |
| Time (weeks) | -0.06 | -0.08, -0.04 | <0.001 |
| NTX-BUP arm X Time | -0.01 | -0.03, 0.03 | 0.767 |

## Appendix D

Predicted average weekly probability of positive urine drug screen (UDS) for methamphetamine in Additive Pharmacotherapy Treatment for methamphetamine disorder (ADAPT-2) randomized controlled trial of naltrexone-bupropion (NTX-BUP) versus placebo.

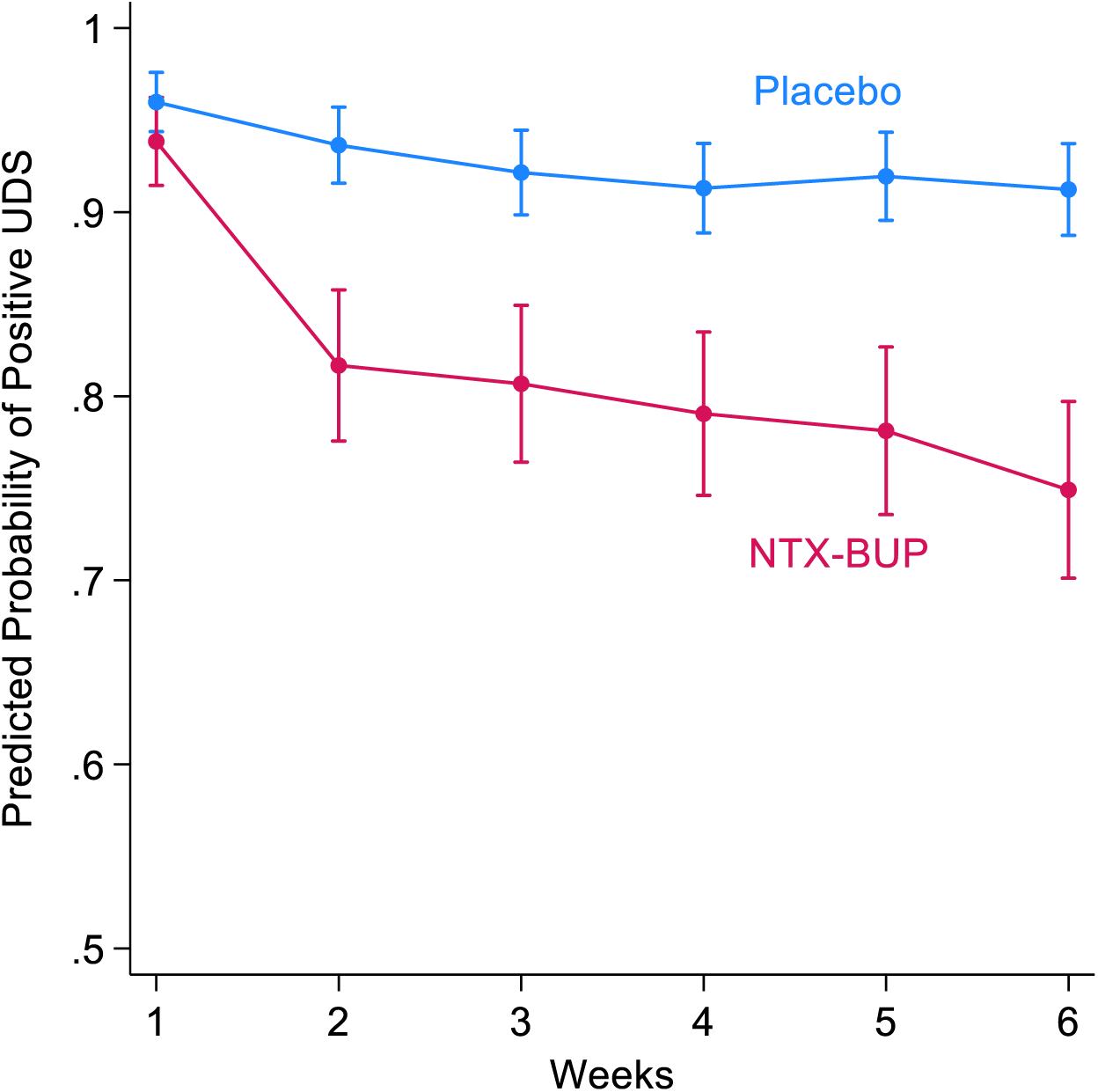

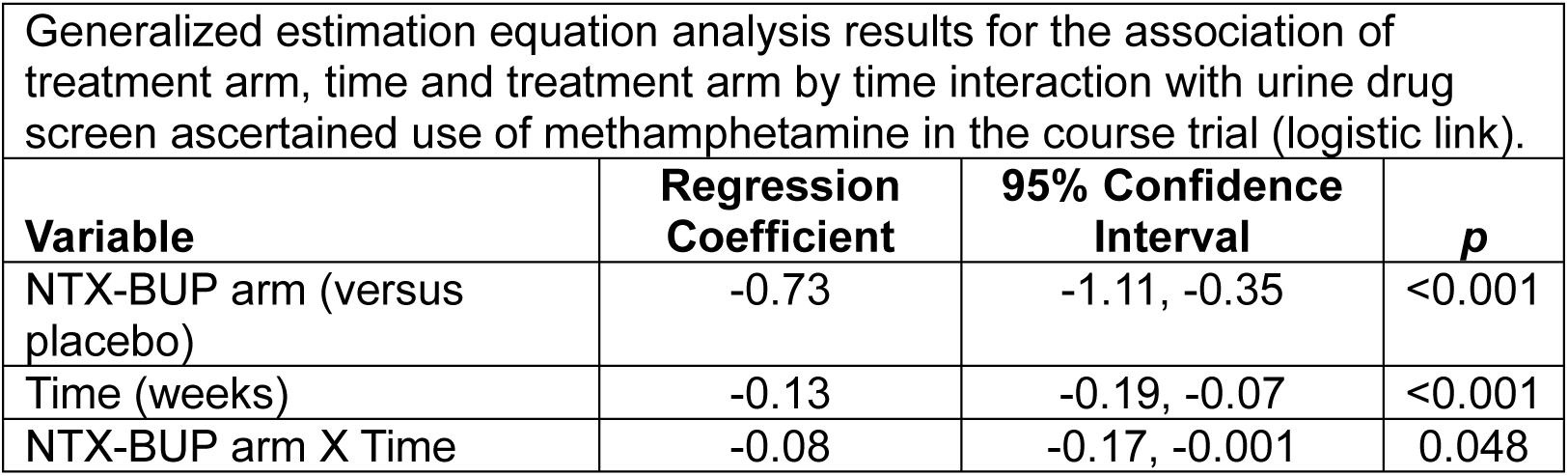

## Appendix E

Predicted average weekly rating of craving in Additive Pharmacotherapy Treatment for methamphetamine disorder (ADAPT-2) randomized controlled trial of naltrexone-bupropion (NTX-BUP) versus placebo.

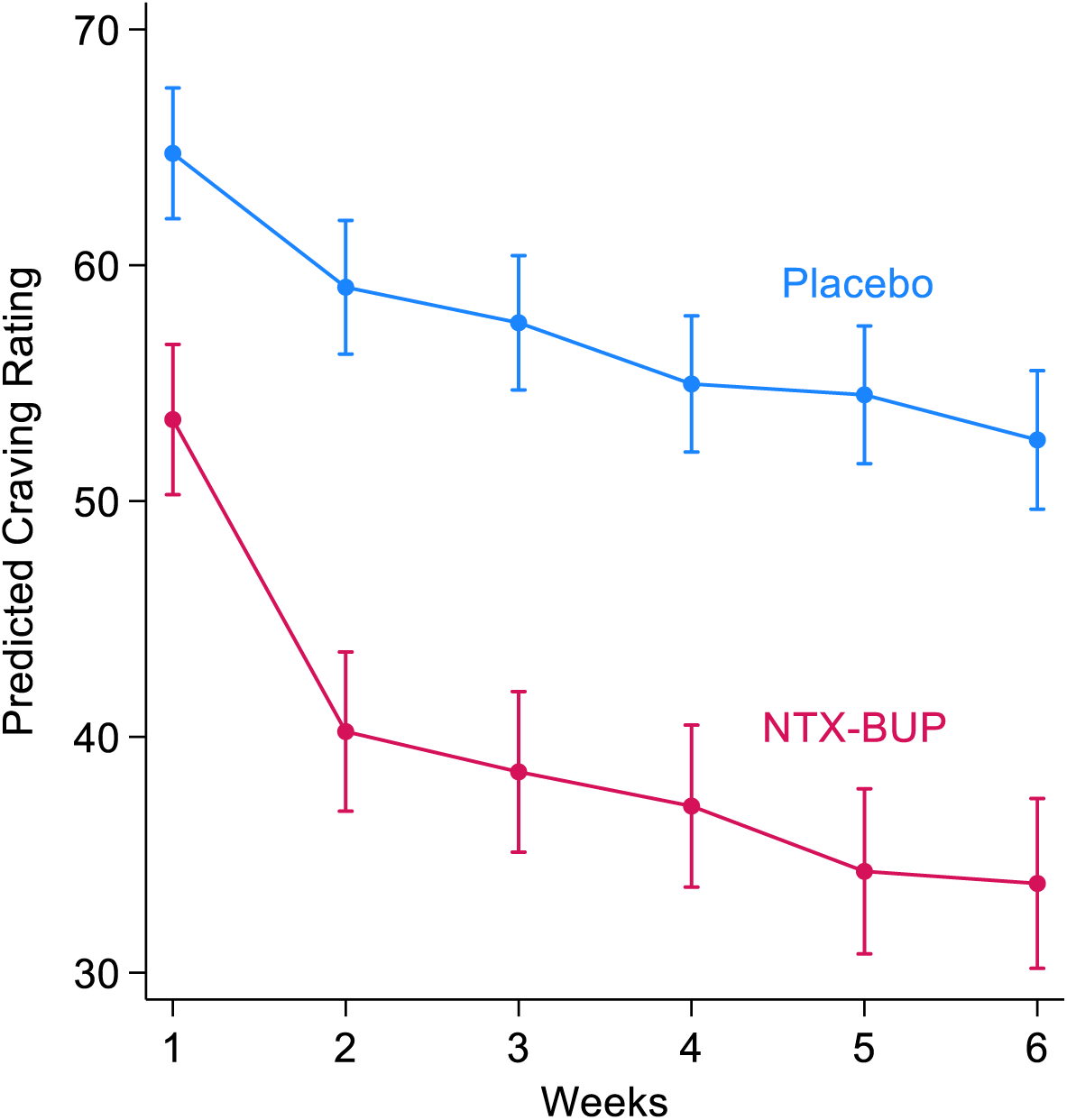

| Generalized estimation equation analysis results for the association of treatment arm, time and treatment arm by time interaction with craving in the course trial (identity link). |  |  |  |
| --- | --- | --- | --- |
| Variable | Regression Coefficient | 95% Confidence Interval | p |
| NTX-BUP arm (versus placebo) | -12.89 | -16.07, -9.70 | <0.001 |
| Time (weeks) | -2.25 | -2.70, -1.80 | <0.001 |
| NTX-BUP arm X Time | -1.32 | -2.09, -0.54 | 0.001 |

## Appendix F

Predicted average weekly rating of depressive symptoms (PHQ-9 score) in Additive Pharmacotherapy Treatment for methamphetamine disorder (ADAPT-2) randomized controlled trial of naltrexone-bupropion (NTX-BUP) versus placebo.

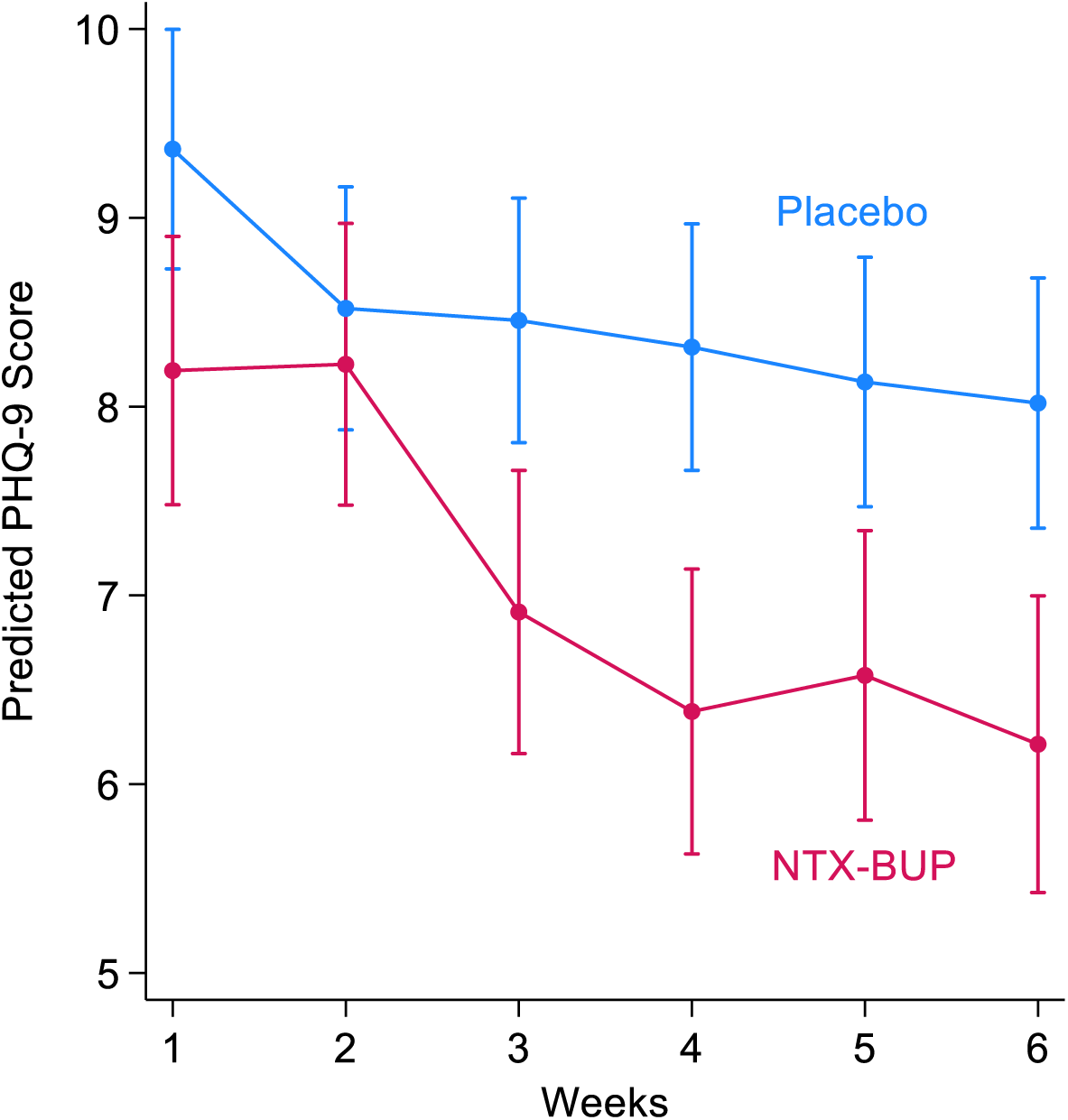

| Variable | Regression Coefficient | 95% Confidence Interval | *p* |
| --- | --- | --- | --- |
| NTX-BUP arm (versus placebo) | -0.66 | -1.31, -0.01 | 0.047 |
| Time (weeks) | -0.24 | -0.33, -0.14 | <0.001 |
| NTX-BUP arm X Time | -0.21 | -0.37, -0.05 | 0.009 |

## Appendix G

Predicted average weekly ratings of impulsivity in Additive Pharmacotherapy Treatment for methamphetamine disorder (ADAPT-2) randomized controlled trial of naltrexone-bupropion (NTX-BUP) versus placebo.

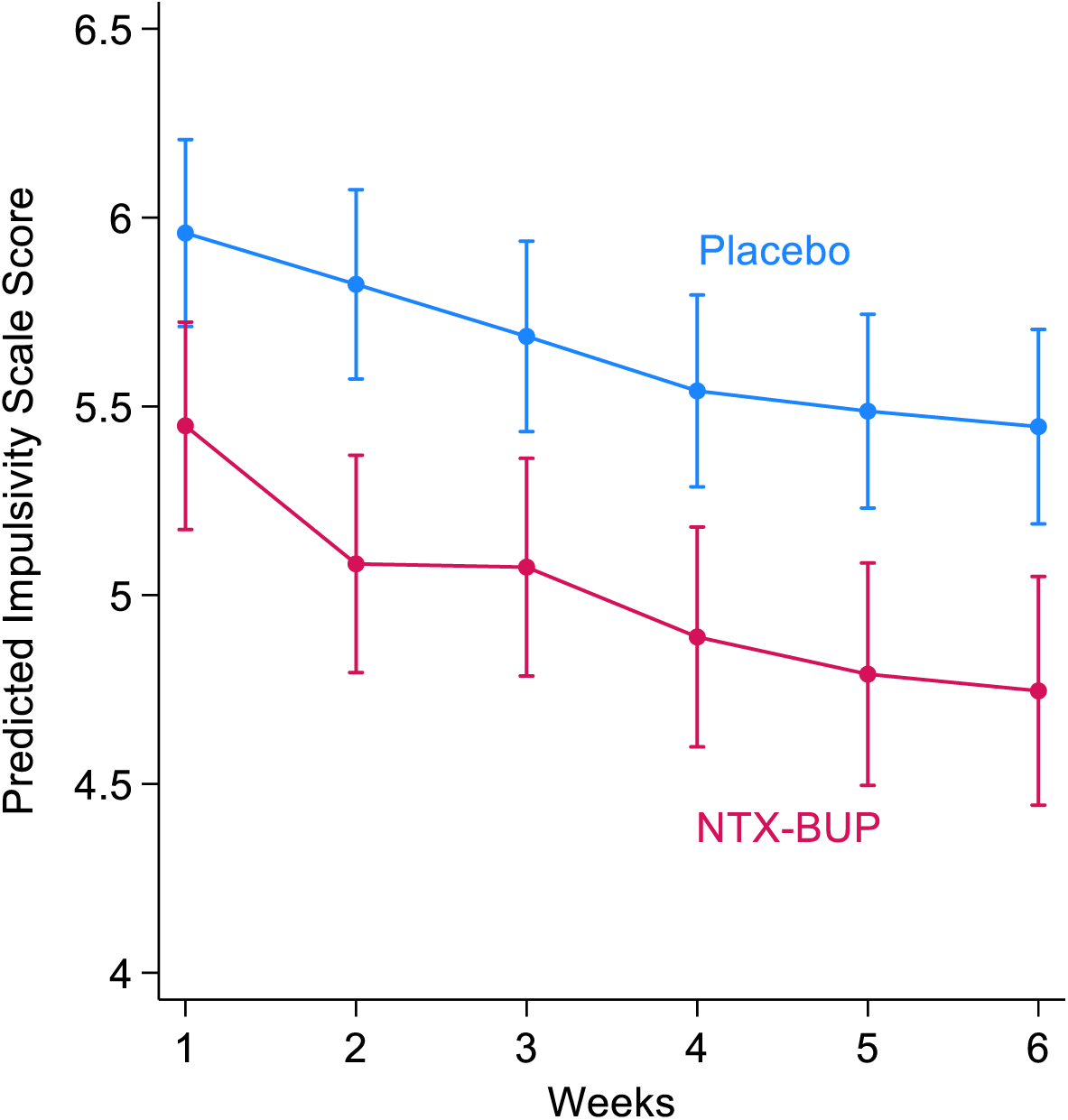

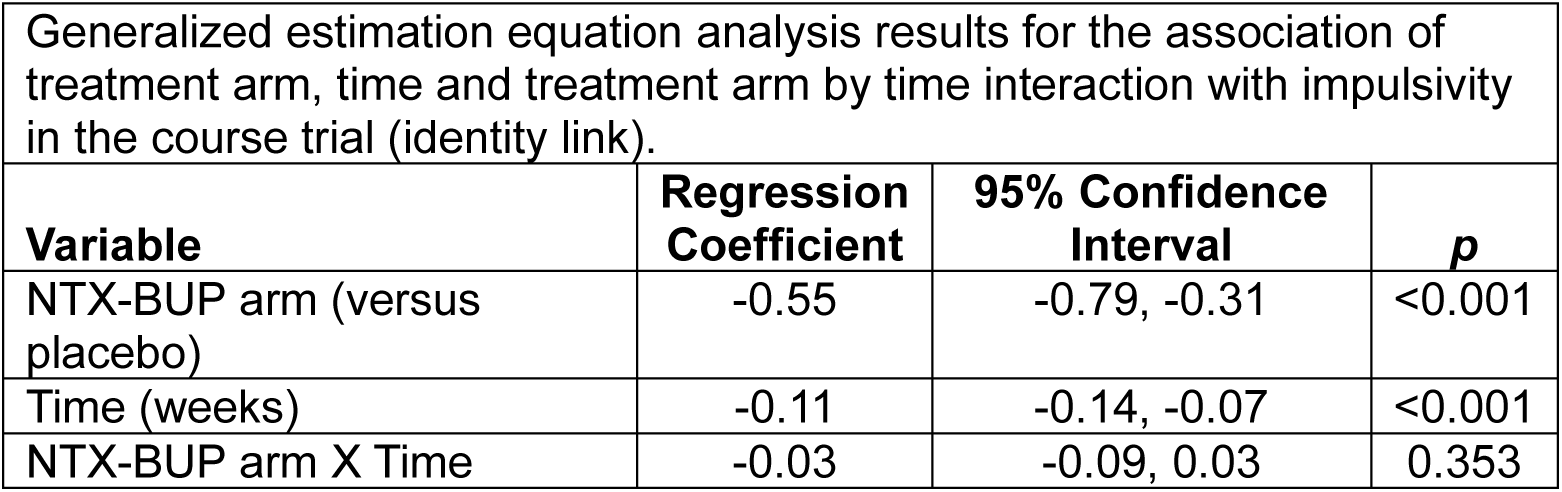

